# Multiple Biomarkers to Predict Major Adverse Cardiovascular Events in Patients With Coronary Chronic Total Occlusions

**DOI:** 10.1101/2023.07.19.23292911

**Authors:** Srikanth Adusumalli, Reza Mohebi, Cian P. McCarthy, Craig A. Megaret, Rhonda F. Rhyne, Farouc A. Jaffer, James L. Januzzi

## Abstract

**Background:** There are limited tools available to predict the long-term prognosis of persons with coronary chronic total occlusions (CTO).

**Objectives:** We evaluated performance of a blood biomarker panel to predict cardiovascular (CV) events in patients with CTO.

**Methods:** From 1251 patients in the CASABLANCA study, 241 participants with a CTO were followed for an average of 4 years for occurrence of major adverse CV events (MACE, CV death, non-fatal myocardial infarction or stroke) and CV death/heart failure (HF) hospitalization. Results of a biomarker panel (kidney injury molecule-1, N-terminal pro-B-type natriuretic peptide, osteopontin, and tissue inhibitor of metalloproteinase-1) from baseline samples were expressed as low-, moderate-, and high-risk.

**Results:** By 4 years, a total of 67 (27.8%) MACE events and 56 (23.2%) CV death/HF hospitalization events occurred. The C-statistic of the panel for MACE through 4 years was 0.79. Considering patients in the low-risk group as a reference, the hazard ratio of MACE by 4 years was 6.65 (95% confidence interval [CI]: 2.98-14.8) and 12.4 (95% CI:5.17-29.6) for the moderate and high-risk groups (both P <0.001). The C-statistic for CVD/HF hospitalization by 4 years was 0.84. Compared to the low-risk score group, the moderate and high-risk groups had hazard ratios of 5.61 (95% CI: 2.33-13.5) and 15.6 (95% CI: 6.18, 39.2; both P value <0.001).

**Conclusion:** A multiple biomarker panel assists in evaluating the risk of adverse outcomes in patients with coronary CTO. These results may have implications for patient care and could have a role for clinical trial enrichment.

**Clinical Trial:** CASABLANCA, ClinicalTrials.gov Identifier: NCT00842868

---

Approximately 1 in 4 patients with obstructive coronary artery disease on coronary angiography have a chronic total occlusion (CTO)^1^, defined as 100 percent stenosis of a coronary artery with Thrombolysis In Myocardial Infarction (TIMI) 0 flow for a duration of at least three months^2^. These common coronary lesions are associated with a worse overall prognosis, with higher rates of major adverse cardiovascular events (MACE)^3–5^. While the symptomatic and prognostic benefit of CTO percutaneous coronary intervention (PCI) is supported by observational studies and registries^6–9^, randomized controlled trial evidence is conflicting regarding the role of CTO PCI in improving patient outcomes^10–15^. Therefore, new approaches to stratify the risk of CTO patients could provide clinical value and inform future randomized trials studying CTO PCI for prognostic benefit.

Despite its potential benefits, CTO PCI has a potentially substantial risk for peri-procedural complications including the risk of coronary artery perforation^6, 16^. Presently the primary indication for CTO PCI is limiting ischemic symptoms, despite prescribed optimal medical treatment. Rather than relying on such subjective assessment, objective measures to stratify risk among individuals with CTO would be useful to further support clinical decision-making. Surprisingly, minimal data is available regarding accurate risk stratification models to predict long-term clinical outcomes and prognosis in CTO patients. An accurate means for risk stratification could aid clinicians in making informed decisions about intensified/personalized medical therapy and inform risk/benefit ratio of CTO revascularization, including CTO PCI.

In the CASABLANCA (Catheter Sampled Blood Archive in Cardiovascular Diseases) study of individuals undergoing coronary angiography we previously described a multi-biomarker panel developed using targeted proteomics and machine learning (HART CVE; Prevencio, Inc, Kirkland WA). This panel, consisting of a weighted algorithm incorporating results from N-terminal pro-B type natriuretic peptide (NT-proBNP), kidney injury molecule-1 (KIM-1), osteopontin, and tissue inhibitor of matrix metalloproteinase-1 (TIMP-1) was highly accurate for predicting incident triple MACE, including non-fatal myocardial infarction (MI), non-fatal stroke, and cardiovascular (CV) death^17^. In the present analysis, we evaluated the performance of this biomarker panel in study participants with one or more CTO. We hypothesized that these novel biomarkers would allow us to risk-stratify patients with CTOs for incident triple MACE. Additionally, as individuals with CTOs often have symptoms of dyspnea and suffer heart failure (HF) events^18^ we also examined the ability of the biomarker panel to predict incident CV death or heart failure (HF) hospitalization.

## Methods

All study procedures were approved by the MassGeneral/Brigham HealthCare Institutional Review Board and conducted in accordance with the Declaration of Helsinki. The design of the CASABLANCA study has been described previously^17^. Briefly, a sample of 1,251 patients undergoing coronary and/or peripheral angiography with or without intervention were prospectively enrolled at the Massachusetts General Hospital in Boston, Massachusetts between 2008 and 2011. Patients were referred for these procedures for numerous reasons, including angiography after acute myocardial infarction (MI), unstable angina pectoris, and HF, as well as for nonacute indications, such as stable chest pain and abnormal stress testing or pre-operatively before heart valve surgery.

After obtaining informed consent, detailed clinical and historical variables and reasons for angiography were recorded at the time of the procedure. Results of coronary angiography (based on visual estimation at the time of the procedure) were recorded; the left main, left anterior descending, left circumflex, and right coronary artery were each considered major coronary arteries, and the highest percent stenosis within each major coronary artery or their branches was recorded. Detailed follow up was then undertaken for cardiovascular (CV), cerebrovascular and renal events and adjudicated using charter definitions. Outcomes assessed for an average of 3.7 years (maximum 8 years) included triple MACE (CV death/non-fatal MI/non-fatal stroke) and CV death/HF hospitalization.

For the purposes of this analysis, all angiograms of study participants with 100% occlusions of a coronary vessel from their case report form were reviewed by the study author (SA). Those with CTO vessels (n=309) were further categorized CTOs into single-vessel, double-vessel, and triple-vessel CTOs. Presence of patent coronary bypass grafts to the CTO was determined. Those with a CTO but with a supplying patent graft (n=68) were excluded from the present analysis, leaving 241 individuals with at least 1 CTO for study analysis (**Figure 1**).

**Figure 1:**
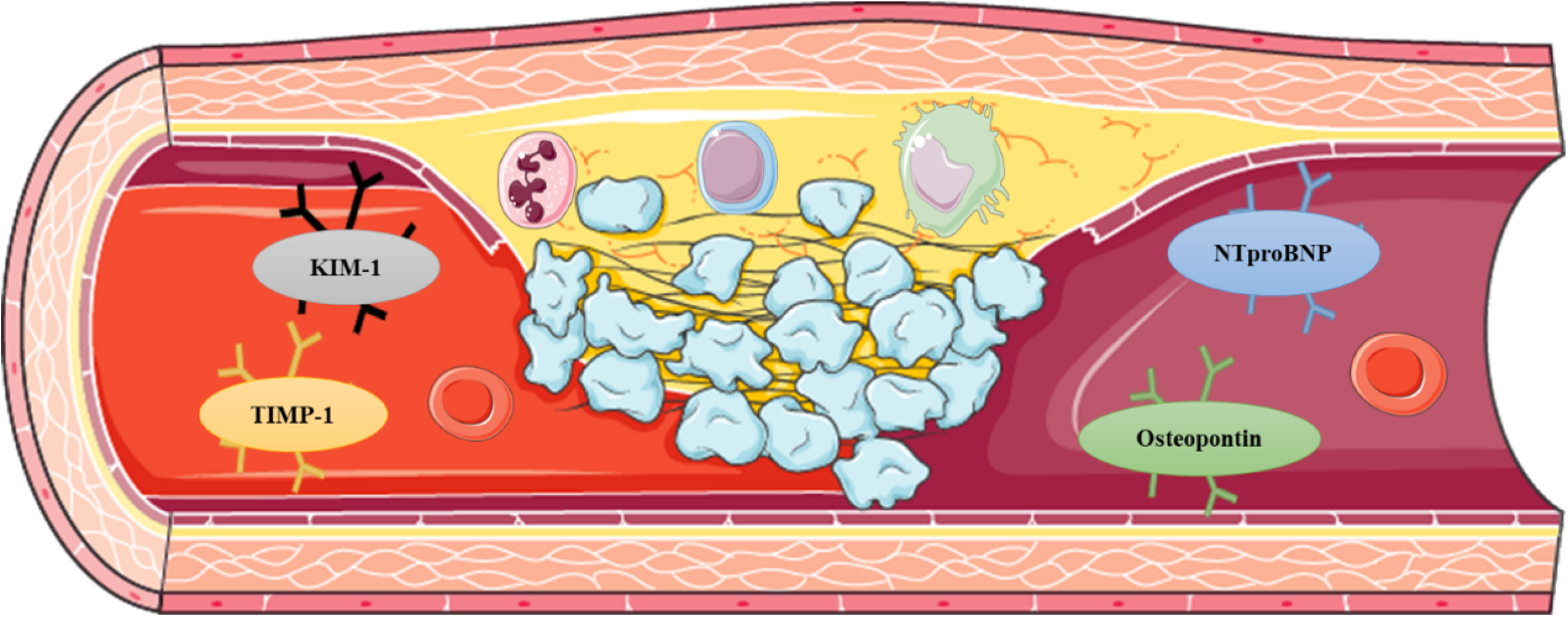
Study flow chart of patients with ≥1 chronic total occlusion (CTO) vessel, excluding those with a patent graft supplying the CTO vessel.

### HART CVE Panel

15 mL of blood was obtained immediately pre- and post-catheterization, processed and frozen at -80 degrees Celsius. Blood was collected into chilled tubes, transported on ice, and centrifuged immediately. Following centrifugation, samples were aliquoted into freezer tubes and frozen immediately at -80 degrees. Analyses are typically performed on aliquots that have not been thawed. In this study, Luminex xMAP technology platform (Luminex Corporation, Austin, Texas) was used to measure concentrations of the 109 proteins listed in **Supplemental Table 1**. The Luminex approach uses multiplexed, microsphere-based assays in a single reaction vessel and is accomplished by assigning each protein-specific assay a recognizable microsphere-based fluorescence signature.

The HART CVE panel (Prevencio, Kirkland WA) was previously developed using an artificial intelligence-leveraged algorithm^19^. After evaluating 109 candidate proteins, a panel of 4 biomarkers (NT-proBNP, KIM-1, osteopontin, and TIMP-1) was derived and used to train a mathematical algorithm, which comprises the HART CVE test panel. This panel results in a continuous result, that is divided into three categories corresponding to low-, medium-, and high-risk levels for CVE. Accordingly, in this analysis study participants were customarily classified into one of those three risk levels.

### Statistical analysis

The distributions of variables were presented using mean (standard deviation), median (interquartile ranges), and count (frequency) as appropriate. Statistical significance of the variables was determined with the chi-square test for trend in proportions (for binary variables) or Spearman’s correlation coefficient test (for continuous variables).

Cox proportional hazard regression was used to calculate the hazard ratio (95% confidence interval) of CVE risk levels for MACE and CV death/HF hospitalization outcomes. The primary analysis of interest was ability of the panel to predict each outcome by 4 years, however outcomes at 1 year, 2 years, and 4 years were all scrutinized. Cox proportional hazard regression assumptions were met using Schoenfeld residual test. Model discrimination and calibration were evaluated with Harrell’s c-statistic and Hosmer-Lemeshow test along with Aikake Information and Bayesian Information Criteria respectively. Statistical significance is defined as a 2-sided P value <0.05. All statistical analyses were performed using the R version 4.3.0 (R Foundation for Statistical Computing, Vienna, Austria. URL: https://www.R-project.org/).

## Results

From the CASABLANCA study, 241 study participants with at least 1 CTO (and without patent bypass graft sources of distal perfusion) were evaluated using the HART CVE panel. Baseline characteristics of study subjects were divided into three groups as low-, medium- and high-risk as detailed in **Table 1**. Medical comorbidities were similar among all risk groups except patients in higher risk group had higher prevalence of pre-existing HF. The median concentrations of KIM-1, NT-proBNP, osteopontin, and TIMP-1 were significantly higher in the high-risk group, as compared to the low and median risk groups.

**Table 1:** Baseline characteristics as a function of HART CVE Score.

| Variable | Low Risk<br>(N=100) | Medium Risk<br>(N=109) | High Risk<br>(N=32) | P-value |
| --- | --- | --- | --- | --- |
| **Demographics** |  |  |  |  |
| Age (Mean/SD) | 62.5 (9.5) | 71.3 (10.1) | 69.5 (12.8) | <0.001 |
| Male sex | 92% | 84.4% | 65.6% | 0.07 |
| Caucasian | 95% | 94.5% | 93.8% | 0.57 |
| **Vital signs (mean/SD)** |  |  |  |  |
| Heart rate (beat/min) | 67.6 (13.3) | 66.4 (12) | 73.9 (14) | 0.14 |
| Systolic BP (mmHg) | 137.7 (19.5) | 141.2 (24.7) | 135.8 (31.1) | 0.59 |
| Diastolic BP (mmHg) | 75.4 (10.4) | 71.4 (12.4) | 70.4 (11.2) | 0.003 |
| **Medical history** |  |  |  |  |
| Smoker | 12.1% | 16% | 3.1% | 0.73 |
| Atrial fibrillation/flutter | 15% | 24.8% | 28.1% | 0.98 |
| Hypertension | 76% | 91.7% | 96.9% | 0.38 |
| Coronary artery disease | 62% | 82.6% | 68.8% | 0.55 |
| Prior MI | 37% | 37.6% | 40.6% | 0.99 |
| Heart failure | 18% | 24.8% | 53.1% | 0.64 |
| Peripheral artery disease | 11% | 33.9% | 56.2% | 0.30 |
| COPD | 13% | 21.1% | 12.5% | 0.78 |
| Diabetes type I/type II | 27% | 40.4% | 53.1% | 0.90 |
| CVA/TIA | 7% | 14.7% | 15.6% | 0.64 |
| CKD | 3% | 20.2% | 53.1% | 0.07 |
| Renal Replacement Therapy | 0% | 0% | 31.2% | 0.32 |
| Prior angioplasty | 11% | 21.1% | 15.6% | 0.20 |
| Prior stent | 32% | 37.6% | 34.4% | 0.46 |
| Prior CABG | 26% | 41.3% | 46.9% | 0.11 |
| History of PCI or other intervention | 30% | 33.9% | 21.9% | 0.44 |
| **Medications** |  |  |  |  |
| ACE-I/ARB | 57% | 71.6% | 62.5% | 0.34 |
| Beta blocker | 77% | 88% | 90.6% | 0.71 |
| Aldosterone antagonist | 3% | 9.3% | 9.4% | 0.38 |
| Loop diuretics | 7% | 22.2% | 59.4% | 0.61 |
| Nitrates | 22% | 32.7% | 34.4% | 0.06 |
| CCB | 22% | 31.2% | 40.6% | 0.39 |
| Statin | 86% | 88% | 75% | 0.29 |
| Aspirin | 87.9% | 86.2% | 71.9% | 0.10 |
| Warfarin | 12% | 18.5% | 21.9% | 0.39 |
| Clopidogrel | 20% | 24.1% | 25% | 0.84 |
| **Echocardiographic results (Mean/SD)** |  |  |  |  |
| LVEF (%) | 57.8 (11.8) | 50.8 (15.4) | 46.2 (19) | 0.002 |
| RSVP | 36.5 (7.1) | 42.1 (10.2) | 48.6 (9.7) | < 0.001 |
| **Stress test results** |  |  |  |  |
| Ischemia on Scan | 90% | 91.9% | 88.9% | 0.37 |
| Ischemia on ECG | 55.9% | 54.8% | 33.3% | 0.78 |
| **Angiographic obstruction results** |  |  |  |  |
| 100% coronary stenosis in 1 vessel | 59.6% | 54.2% | 65.6% | 0.56 |
| 100% coronary stenosis in 2 vessels | 27.3% | 31.8% | 18.8% | 0.60 |
| 100% coronary stenosis in 3+ vessels | 12.1% | 14% | 15.6% | 0.23 |
| **Lab measures** |  |  |  |  |
| Sodium (mean/SD) | 139.8 (2.5) | 139 (2.9) | 137.8 (4.4) | 0.03 |
| Blood urea nitrogen (median Q1, Q3) | 17 (14, 19.2) | 20 (15, 28) | 28 (23, 46) | <0.001 |
| Creatinine (median Q1, Q3) | 1 (0.9, 1.2) | 1.2 (1, 1.5) | 1.7 (1.4, 4.6) | <0.001 |
| eGFR (median Q1, Q3) | 109.5 (102.7, 117.6) | 82.9 (66.2, 102.8) | 39.6 (10.9, 56.2) | <0.001 |
| Total cholesterol (median Q1, Q3) | 151.8 (42.3) | 134.9 (36.9) | 134.2 (56) | <0.001 |
| LDL cholesterol (median Q1, Q3) | 84 (35.6) | 68.7 (25.5) | 70.5 (48.1) | <0.001 |
| Glycohemoglobin (median Q1, Q3) | 6.2 (5.7, 6.6) | 6.2 (5.6, 6.9) | 6.7 (6.3, 7.6) | 0.23 |
| Glucose (median Q1, Q3) | 98.5 (89, 108.2) | 100 (92, 142) | 120 (95.8, 143.8) | 0.006 |
| Hemoglobin (Mean/SD) | 14 (1.5) | 13 (1.7) | 11.7 (1.6) | <0.001 |
| **Prognostic Panel Biomarkers** |  |  |  |  |
| KIM-1, ng/mL (median Q1, Q3) | 0.015 (0.015, 0.034) | 0.059 (0.045, 0.09) | 0.15 (0.082, 0.278) | <0.001 |
| NT-proBNP, pg/mL (median Q1, Q3) | 645 (251, 1170) | 2240 (1220, 4430) | 15975 (8078, 15975) | <0.001 |
| Osteopontin, ng/mL (median Q1, Q3) | 19 (14.8, 27.2) | 33 (25, 44) | 81.5 (60.8, 105) | <0.001 |
| TIMP-1, ng/mL (median Q1, Q3) | 63.5 (55, 71) | 78 (67, 96) | 124 (110.2, 172.2) | <0.001 |
| ACE-I – angiotensin converting enzyme inhibitor, ARB – angiotensin receptor blocker, CCB -calcium channel blockers, BP- blood pressure, CVA – cerebrovascular accident, CABG – coronary artery bypass grafting, CKD – chronic kidney disease, ECG – electrocardiogram, eGFR – estimated glomerular filtration rate, LDL – low density lipoproteins, LVEF – Left ventricular ejection fraction, MI- myocardial infarction, PCI – Percutaneous coronary intervention, RVSP – Right ventricular systolic pressure, TIMP-1 – tissue inhibitor of matrix metalloproteinase, TIA – Transient ischemic attack. |  |  |  |  |

### Distribution of CVE panel by the extent of CTOs

The coronary risk status of participants was then considered as a function of their HART CVE result, expressed as low-, medium-, or high-risk. As displayed in **Table 2**, study participants with a high-risk HART CVE result had a greater number of vessels with >70% vessel stenosis compared to those with a low-risk result (Mean number of vessels: 1.57 vs. 1.13, p-value: 0.02), while the distribution of 1-, 2-, or 3-vessel CTO was comparable across HART CVE groupings.

**Table 2:** Distribution of CVE Panel by the extent of CTO.

| **Variable** | **All** | **CVE score**<br>**low** | **CVE score**<br>**moderate** | **CVE score**<br>**high** | **P-value across**<br>**low/moderate/high** |
| --- | --- | --- | --- | --- | --- |
| Mean (SD) number of vessels with >70% stenosis | 1.248 (1.429) | 1.132 (1.316) | 1.281 (1.489) | 1.569 (1.557) | 0.02 |
| Single vessel CTO | 138 (11.1%) | 59 (11%) | 58 (10.3%) | 21 (15.3%) | 0.37 |
| 2 vessel CTO | 67 (5.4%) | 27 (5%) | 34 (6%) | 6 (4.4%) | 0.90 |
| 3 vessel CTO | 32 (2.6%) | 12 (2.2%) | 15 (2.7%) | 5 (3.6%) | 0.37 |
| CTO – Chronic total occlusions; CVE – Cardiovascular events |  |  |  |  |  |

### Performance of HART CVE Panel for triple MACE and CV death/HF hospitalization

There were 40 (16.5%), 51 (21.1%) and 67 (27.8%) MACE events at 1, 2 and 4-year follow up respectively and 35 (14.5%), 42 (17.4%), 56 (23.2%) CV death/ HF hospitalisation events at 1, 2 & 4 years follow up respectively.

The C-statistic of the model for predicting 4-year triple MACE was 0.79 (0.72, 0.85). As shown in **Table 3**, compared to the referent low-risk group, by one year, those with a moderate-risk HART CVE result had a hazard ratio (HR) with 95% confidence interval (CI) for MACE at 1 year of 5.32 (95% CI: 1.98, 14.3; P <0.001), while a high-risk HART CVE result had an associated HR of 14.7 (95% CI: 5.28, 40.8; P <0.001). Similar graded results for triple MACE were seen associated with moderate- and high-risk CVE scores at 2 years (moderate score HR: 5.46, 95% CI 1.84, 8.77; high score HR:14.1, 95% CI: 5.59, 35.5; both P <0.001) and at the 4-year time point (moderate score HR 6.65, 95% CI 2.98, 14.8; high score HR: 12.4, 95% CI: 5.17, 29.6; both P <0.001). The model was well-calibrated **(Table 4).**

**Table 3:** Performance of HART CVE panel for triple MACE across study time points.

| Time frame | CVE risk category | HR | 95% CI | P-value |
| --- | --- | --- | --- | --- |
| 1 Year |  |  |  |  |
|  | Low risk | referent | referent | referent |
|  | Moderate risk | 5.32 | 1.98, 14.3 | 0.001 |
|  | High risk | 14.67 | 5.28, 40.8 | <0.001 |
| 2 Year |  |  |  |  |
|  | Low risk | referent | referent | referent |
|  | Moderate risk | 5.46 | 1.84, 8.77 | <0.001 |
|  | High risk | 14.08 | 5.59, 35.5 | <0.001 |
| 4 Year |  |  |  |  |
|  | Low risk | referent | referent | referent |
|  | Moderate risk | 6.65 | 2.98, 14.8 | <0.001 |
|  | High risk | 12.37 | 5.17, 29.6 | <0.001 |
| CVE – Cardiovascular events, MACE – Major adverse cardiovascular events |  |  |  |  |

**Table 4:** Calibration for triple MACE.

| **Features** | **AIC** | **BIC** | **H-L P-value** |
| --- | --- | --- | --- |
| NT-proBNP | 187.159 | 194.1286 | 0.22 |
| NT-proBNP + KIM-1 | 188.2956 | 198.75 | 0.99 |
| NT-proBNP + KIM-1 + OPN | 188.0387 | 201.9779 | 1.0 |
| NT-proBNP + KIM-1 + OPN + TIMP-1 | 185.344 | 202.768 | 0.39 |
| AIC = Akaike information criterion, BIC = Bayesian information criterion, H-L = Hosmer–Lemeshow test, KIM-1 – kidney injury molecule-1, MACE – Major adverse cardiovascular events, NT-proBNP – N-terminal pro-B-type natriuretic peptide, OPN-osteopontin, TIMP-1 - tissue inhibitor of metalloproteinase-1 |  |  |  |

Associations between HART CVE score and risk of MACE as a function of number of CTO vessels are detailed in **Supplemental Table 2**. This shows generally higher risk associated with more CTO vessels, particularly with higher HART CVE scores. Across each category of CTO extent, those with low HART CVE scores had similar HR across 1-, 2-, or 3-vessel CTO groups. These consistent results across 1-3 vessel CTO patients indicates robustness of the biomarker panel across a spectrum of CTO patients.

Relevant to the CV death/HF hospitalization outcome the C-statistic of the model for predicting 4-year CV death/HF hospitalization was 0.84 (0.78, 0.90). **Table 5** displays the performance of HART CVE panel for CV death/HF hospitalization across time points. This shows, comparable to the performance of the HART CVE panel to predict triple MACE, graded risk across low-, medium- and high-risk scores was seen for CV death/HF hospitalization at each time point examined. For example, compared to the referent (the low-risk score group), the high-risk group had a hazard ratio of 26.1 (95% CI: 7.55, 90.2) at 1 year, 16.8 (95% CI: 6.11, 46.2) at 2 years and 15.6 (95% CI: 6.18, 39.2) at 4 years (all P value <0.001). In addition, the model’s calibration was excellent **(Table 6).**

**Table 5:** Performance of HART CVE Panel for CV death/HF hospitalization across study time points.

| Time frame | CVE risk category | HR | 95% CI | P-value |
| --- | --- | --- | --- | --- |
| 1 Year |  |  |  |  |
|  | Low-risk | referent | referent | referent |
|  | Moderate-risk | 6.56 | (1.89, 22.7) | 0.003 |
|  | High-risk | 26.1 | (7.55, 90.2) | <0.001 |
| 2 Year |  |  |  |  |
|  | Low-risk | referent | Referent | referent |
|  | Moderate-risk | 4.96 | (1.86, 13.2) | 0.001 |
|  | High-risk | 16.8 | (6.11, 46.2) | <0.001 |
| 4 Year |  |  |  |  |
|  | Low-risk | referent | Referent | referent |
|  | Moderate-risk | 5.61 | (2.33, 13.5) | <0.001 |
|  | High-risk | 15.6 | (6.18, 39.2) | <0.001 |
| CVE – Cardiovascular events |  |  |  |  |

**Table 6:** Calibration for CV death/HF hospitalization.

| **Features** | **AIC** | **BIC** | **H-L P-value** |
| --- | --- | --- | --- |
| NT-proBNP | 160.9016 | 167.8712 | 0.67 |
| NT-proBNP + KIM-1 | 162.2871 | 172.7415 | 0.94 |
| NT-proBNP + KIM-1 + OPN | 156.0276 | 169.9668 | 0.42 |
| NT-proBNP + KIM-1 + OPN + TIMP-1 | 156.5591 | 173.9831 | 0.21 |
| AIC = Akaike information criterion, BIC = Bayesian information criterion, H-L = Hosmer–Lemeshow test, KIM-1 – kidney injury molecule-1, MACE – Major adverse cardiovascular events, NT-proBNP – N-terminal pro-B-type natriuretic peptide, OPN- osteopontin, TIMP-1 - tissue inhibitor of metalloproteinase-1 |  |  |  |

As depicted in **Figure 2 and 3** the time to first triple MACE or CV death/HF hospitalization was shortest for study participants with a CTO and a high-risk HART CVE classification.

**Figure 2:**
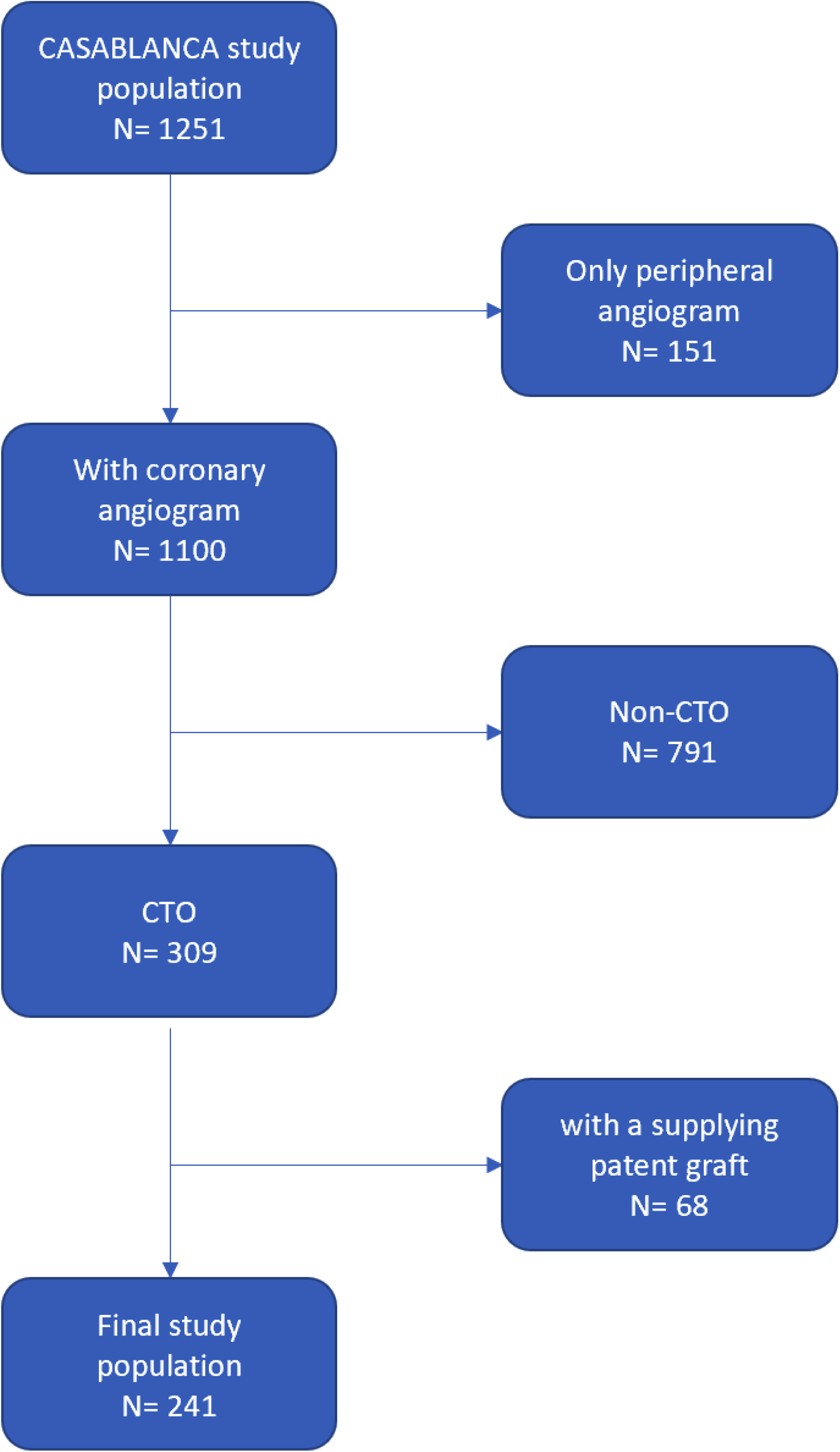
Cumulative event curves over a follow up period of 4 years in patients with ≥1 coronary chronic total occlusion (CTO) vessels categorized by HART CVE risk group and subsequent major adverse cardiovascular events (MACE) rates. Subjects in the high-risk HART CVE group had considerably higher event rates from enrollment to 4 years.

**Figure 3:**
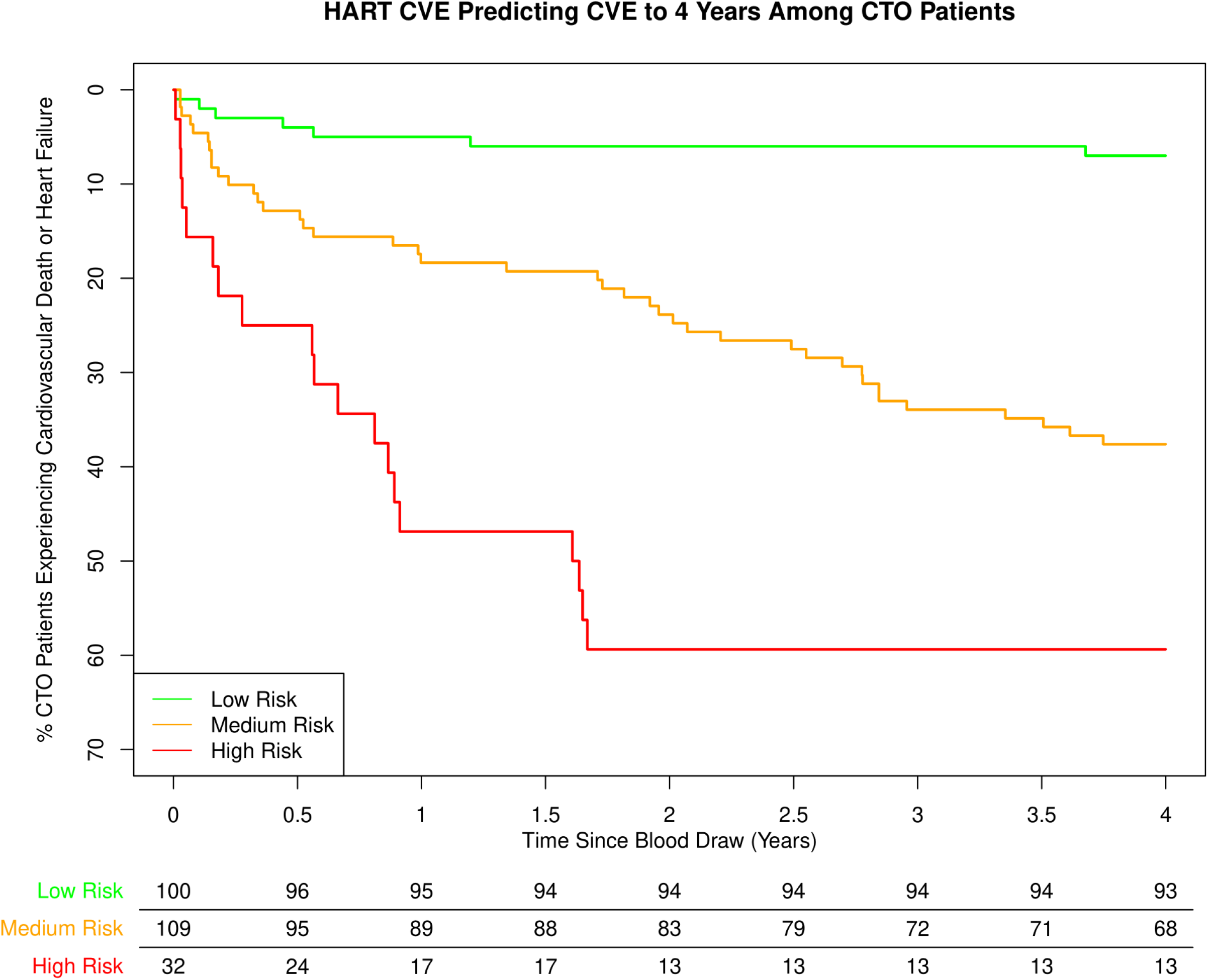
Cumulative event curves over 4-year follow up in patients with coronary ≥1 CTO vessels categorized by HART CVE risk group and subsequent cardiovascular death or heart failure rates. Subjects in the high-risk HART CVE group had considerably higher event rates from enrollment to 4 years.

## Discussion

Among CASABLANCA study patients with CTO lesion that was not bypassed with a patent distal bypass graft, a multimarker weighted panel comprised of four biomarker proteins (NT-proBNP, KIM-1, Osteopontin and TIMP-1) accurately risk stratified CTO patients into low-, medium- and high-risk groups across a very broad range of risk for both triple MACE as well as CV death/HF hospitalization. The HART CVE panel provided excellent discrimination for predicting such risk throughout the 4-year follow-up period. By 4 years, compared to a referent low-risk population, the high-risk group had a greater than 12-fold higher MACE risk during the 4 years of follow-up and nearly 16-fold higher risk for CV death/HF hospitalization during that same period. Even though patients with numerically more CTO vessels had higher risk for adverse outcome, even in this context the biomarker panel remained able to stratify risk for MACE outcomes. The ability to stratify baseline risk in patients with CTOs remains unexplored, so to our knowledge, this study is the first to describe the role of biomarker testing for this indication.

The four biomarkers included in the HART CVE panel reflect various aspects of pathophysiology among patients with CTOs (**Central Illustration**). NT-proBNP, which is released due to LV wall stretch secondary to LV dysfunction^20^ and myocardial ischemia^21^, has also been associated with severity^22, 23^ and long-term prognosis of patients with stable CAD^24–26^. KIM-1 is associated with progressive renal disease^27–29^ but is also elevated in proportion to the presence and severity of CAD^30^. Osteopontin regulates myofibroblast differentiation and production of type I collagen production, is upregulated following myocardial infarction, and associates with cardiac fibrosis^31^. Lastly, TIMP-1 is involved in the degradation of extracellular matrix proteins. Increased expression of TIMP-1 occurs following chronic pressure overload in myocardium, is associated with cardiac fibrosis, and may be linked to coronary plaque instability^32^.

While abundant reports are present regarding models to predict in-hospital/short-term outcomes following CTO PCI, few data exist regarding the use of pre-PCI variables for predicting longer-term clinical risk among individuals with CTO. The results of this study suggest that a validated biomarker panel can stratify risk accurately in individuals with coronary CTO, even among those with numerically more CTO vessels. Arguably, given the risk/benefit trade-off of CTO PCI, having a tool to judge risk for short, medium, and longer-term risk for MACE or HF events may inform personalized and intensified medication administration and follow-up frequency, and may similarly dictate a threshold for proceeding with CTO PCI. For example, among those with a low-risk HART CVE score, regardless of the number of CTO vessels, the MACE risk was low, which might suggest a continued medical therapy strategy, while in patients with similar number of CTO vessels, a higher risk score consistently predicted greater MACE events at 1, 2, and 4 years; such individuals, within the context of shared decision making, might be more reasonably considered for earlier attempt at CTO revascularization for improved prognosis, independent of symptom status.

Another potential use of the HART CVE score could be to enrich clinical trials with CTO patients more likely to experience MACE. For example, patients with moderate or higher HART CVE scores could potentially be part of future randomized control trials investigating the benefits of CTO PCI on outcomes such as MACE and CV death/HF hospitalization providing utilities for sufficient powering of such studies. More precisely, given the event rates in those with the most elevated HART CVE results (60% at 4 years, compared to a typical 4-year event of 25% in a post-ACS population^33^), clinical trials evaluating CTO PCI for improved prognosis could enroll more efficiently if utilizing a pre-stratified population expected to have higher CV event rates based on an elevated HART CVE test. Whether PCI of a CTO as part of a revascularization strategy can reduce the elevated risk seen in those with a higher risk HART CVE panel result remains speculative, but this study—leveraging results from a novel biomarker panel—represents an important first step that creates further possibilities.

Besides triple MACE, in this study we also chose to examine rates of incident CV death/HF hospitalization as an outcome measure, as HF symptoms are common among persons with CTOs^34, 35^ and successful CTO PCI is often associated with dyspnea improvement^34^. The high HF event rate in this study is of note as is the fact that a panel that leverages concentration of NT-proBNP was comparably discriminatory for HF events to 4 years from study enrolment.

### Study limitations

Although the study was performed with a careful methodologic approach, limitations to our study exist. First, the CASABLANCA study participants were predominantly male and Caucasian with a smaller representation of patients with traditional risk factors for CAD such as smoking (12%) and diabetes mellitus (36%). Second, the number of patients from which we derived and validated our findings was relatively small. Third, we only measured biomarkers at a single point in time, which may not reflect levels at future time periods. The study results require further validation and testing in a randomized controlled study. Fourth, the effects of revascularization on MACE during follow-up were not adjusted; salutary effect of revascularization would be expected to undermine the results of the HART CVE test, which was not observed. Finally, whether aggressive treatment strategies such as CTO PCI in patients with high-risk score will modify their risk status and whether that will reflect on long-term outcomes remains unknown but is a testable hypothesis.

In conclusion, a multi-biomarker panel developed using targeted proteomics and machine learning was highly accurate for the risk stratification of patients with CTOs. The panel was able to accurately predict both incident triple MACE as well as CV death/HF hospitalization over a 4-year period. The potential advantages of using a biomarker leveraged approach for decision-making in CTO patients include its use for guiding personalized medical therapy and surveillance, and to help select patients that might benefit prognostically from CTO PCI.

### Clinical Perspective

- **Competency in medical knowledge:** Patients with a coronary chronic total occlusions (CTO) experience elevated MACE rates, but the role of CTO percutaneous coronary intervention (PCI) in reducing MACE is unclear and the procedures itself carries risk. A risk stratification tool for long-term risk CTO patients is therefore needed.
- **Translational outlook 1:** A multi-biomarker panel from CASABLANCA study has previously shown predict MACE.
- **Translational outlook 2:** The HART CVE panel was used to risk stratify CTO patients in to low-, medium- and high-risk groups. Using the low-risk group as reference, the high-risk group had considerably higher incidence of triple MACE, with a 60% incidence of cardiovascular death, nonfatal myocardial infarction, and nonfatal stroke, as well as CV death/HF hospitalization at 4 years.
- **Translational outlook 3:** Using this multi-biomarker panel, physicians may be able to risk stratify and select high-risk CTO patients for enhanced secondary CAD prevention and surveillance, and enrichment for CTO PCI randomized trials.

## Data Availability

All the data for the manuscript is available

---

**Central illustration:**
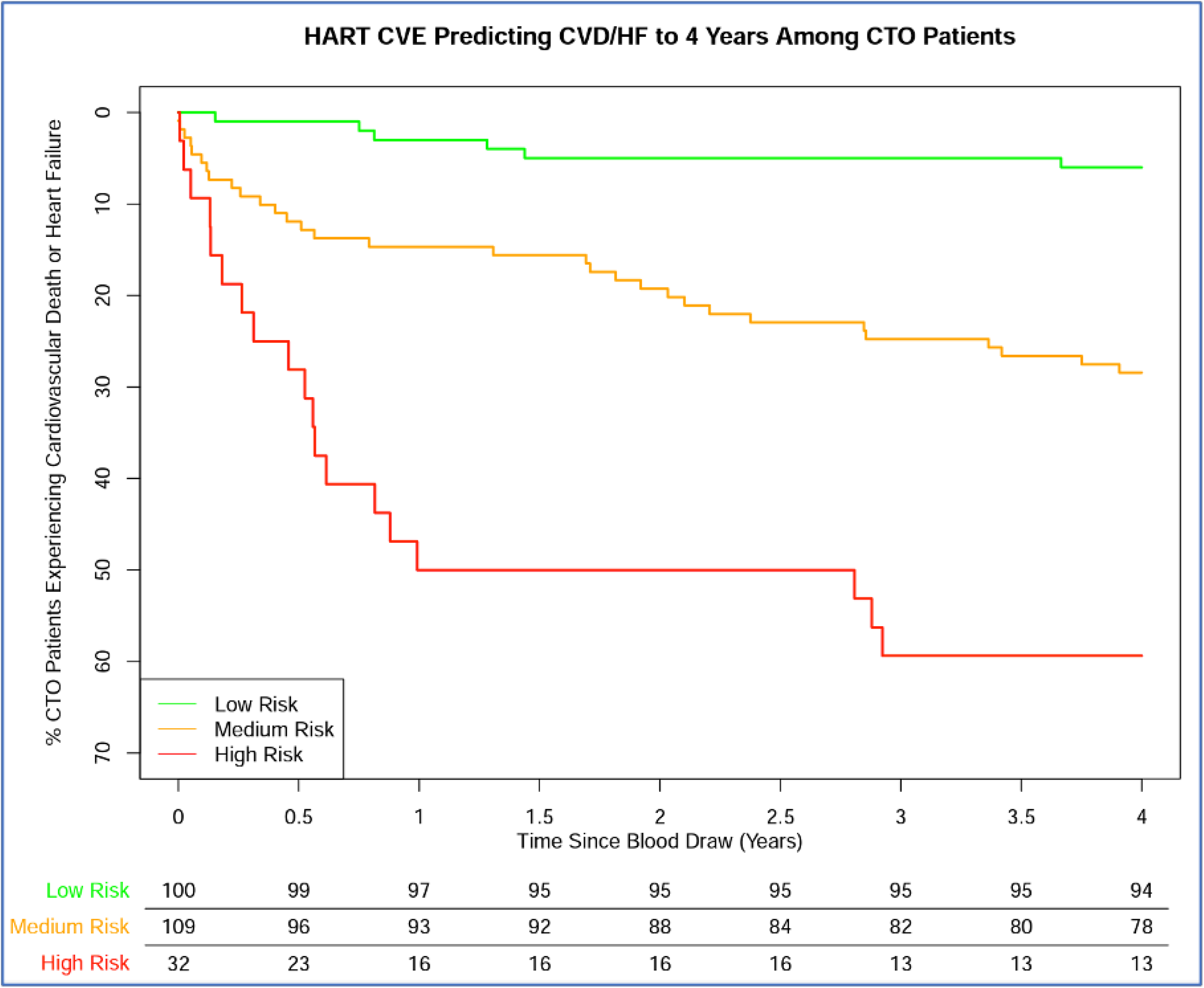
Patients with ≥1 CTO vessel may experience a poorer prognosis stratified by a biomarker panel level (HART CVE) that includes kidney injury molecule-1, N-terminal pro-B-type natriuretic peptide, osteopontin, and tissue inhibitor of metalloproteinase-1. Mechanistically, this finding might reflect progression of higher risk plaques and/or myocardial demand ischemia/infarction, driving an increased risk of major adverse cardiovascular events.

